# Replication of a neuroimaging biomarker for striatal dysfunction in psychosis

**DOI:** 10.1101/2023.07.17.23292779

**Authors:** Jose M Rubio, Todd Lencz, Hengyi Cao, Nina Kraguljac, Elvisha Dhamala, Philipp Homan, Guillermo Horga, Deepak K. Sarpal, Miklos Argyelan, Juan Gallego, John Cholewa, Anita Barber, John Kane, Anil Malhotra

## Abstract

To bring biomarkers closer to clinical application, they should be generalizable, reliable, and maintain performance within the constraints of routine clinical conditions. The functional striatal abnormalities (FSA), is among the most advanced neuroimaging biomarkers in schizophrenia, trained to discriminate diagnosis, with post-hoc analyses indicating prognostic properties. Here, we attempt to replicate its diagnostic capabilities measured by the area under the curve (AUC) in receiver operator characteristic curves discriminating individuals with psychosis (n=101) from healthy controls (n=51) in the Human Connectome Project for Early Psychosis. We also measured the test-retest (run 1 vs 2) and phase encoding direction (i.e., AP vs PA) reliability with intraclass correlation coefficients (ICC). Additionally, we measured effects of scan length on classification accuracy (i.e., AUCs) and reliability (i.e., ICCs). Finally, we tested the prognostic capability of the FSA by the correlation between baseline scores and symptom improvement over 12 weeks of antipsychotic treatment in a separate cohort (n=97). Similar analyses were conducted for the Yeo networks intrinsic connectivity as a reference. The FSA had good/excellent diagnostic discrimination (AUC=75.4%, 95%CI=67.0%-83.3%; in non-affective psychosis AUC=80.5%, 95%CI=72.1-88.0%, and in affective psychosis AUC=58.7%, 95%CI=44.2-72.0%). Test-retest reliability ranged between ICC=0.48 (95%CI=0.35-0.59) and ICC=0.22 (95%CI=0.06-0.36), which was comparable to that of networks intrinsic connectivity. Phase encoding direction reliability for the FSA was ICC=0.51 (95%CI=0.42-0.59), generally lower than for networks intrinsic connectivity. By increasing scan length from 2 to 10 minutes, diagnostic classification of the FSA increased from AUC=71.7% (95%CI=63.1%-80.3%) to 75.4% (95%CI=67.0%-83.3%) and phase encoding direction reliability from ICC=0.29 (95%CI=0.14-0.43) to ICC=0.51 (95%CI=0.42-0.59). FSA scores did not correlate with symptom improvement. These results reassure that the FSA is a generalizable diagnostic – but not prognostic – biomarker. Given the replicable results of the FSA as a diagnostic biomarker trained on case-control datasets, next the development of prognostic biomarkers should be on treatment-response data.

---

Personalized medicine aims to use biomarkers to match individuals with their most appropriate interventions^1^. This is particularly relevant in psychiatry, since most often treatment choice is determined by trial and error, which is associated with treatment disengagement and greater chance of suboptimal outcomes^2^. For personalized medicine to deliver on its promise of improving clinical outcomes, biomarker development should fulfill several criteria. Akin to the phases of drug discovery, these consecutive steps are: identifying a biological measure fit for the clinical paradigm of interest; demonstrating that it scales with the clinical measure of interest independent of potential confounding (internal validity); showing out of sample performance (external validity); and finally proving clinical utility^3^.

In a recent review covering the state of the field in biomarker development^4^, we identified the Functional Striatal Abnormalities (FSA)^5^ index among the most developed in schizophrenia. FSA is a data-driven measure informed by previous evidence on the role of striatal function in schizophrenia. For its development, investigators used functional magnetic resonance images (fMRI) acquired from seven independent scanners (n = 1,100), which were used to derive intra- and extra-striatal functional connectivity and striatal fractional amplitude of low-frequency fluctuations (fALFF) images. Features from these three sources were selected using a support vector machine (SVM) classifier trained to discriminate between patients with non-affective psychosis (n=560) and controls (n=540), and the FSA score value was defined as the distance in the SVM feature space to the separating hyperplane. Polarity was defined so that positive values were predictive for healthy controls. The authors found accuracy of 80.4%, sensitivity of 79.3%, and specificity of 81.5% based on leave-one-site-out cross validation. Despite not having been developed as a predictive biomarker, post-hoc analyses showed that in two separate cohorts for which there were data on treatment response, baseline FSA score was significantly correlated with symptom reduction (r=0.62, 0.42; p<0.01 and p<0.01 respectively), showing also promise as a biomarker of treatment response.

While these data are encouraging, the ability to move biomarkers towards clinical practice is contingent on independent confirmations of external validity^3^. The FSA’s performance was tested by leave-one-site-out cross-validation^5^, but validation of these results by independent groups under generalizable conditions (i.e., different scanners, imaging protocols, or participant characteristics) is still necessary^6^. Furthermore, in addition to testing the accuracy of predictions, several additional requirements for an effective biomarker include characterization of reliability; examination of potential confounding effects by clinical and demographic variables; testing sensitivity to effects of imaging acquisition and analysis parameters. Biomarker stability in the face of these potential confounds is critical to plan subsequent experiments geared towards demonstrating clinical utility.

Here, we aim to advance this line of research on biomarker development in schizophrenia by testing whether the classification performance of the FSA replicates in the Human Connectome Project for Early Psychosis^7^, a new publicly available dataset with different demographic characteristics, acquisition parameters and scanner type to the one in which the FSA was developed. Furthermore, we aim to study parameters that affect the validity and reliability of this biomarker. Specifically, we measured the effects of relevant confounding on FSA values, calculated its test-retest (i.e., run 1 vs run 2) and phase encoding direction reliability (i.e., FSA generated from PA scan vs from AP scan). In addition, we repeated these analyses for intrinsic connectivity for the Yeo networks, which have been well characterized ^8^.

Also, we concatenated scan runs and resliced them to obtain scans of increasing lengths from which we obtained FSA and network intrinsic connectivity to study the effects of increasing scan duration on accuracy of classification and reliability. Finally, since the Human Connectome Project for Early Psychosis^7^ does not have data on treatment response, to replicate the post-hoc finding of the correlation between treatment response and baseline FSA scores, we tested this on a separate cohort of individuals with first episode psychosis who were treated with 12 weeks of antipsychotics (Figure 1).

**Figure 1.**
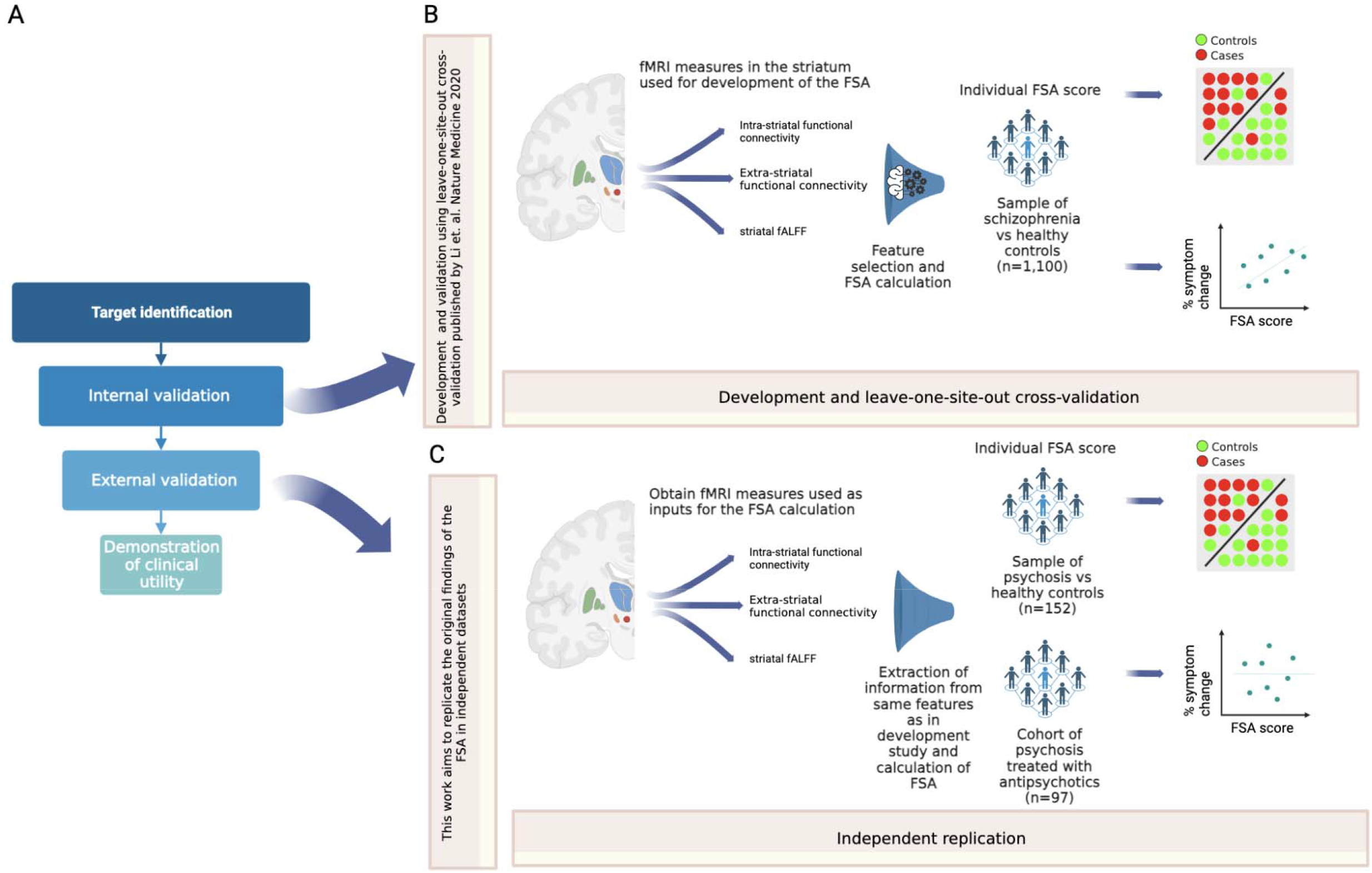
Overall study in context. **Footnote:** A: Framework for the development of biomarkers, as in Abi-Dargham and Horga. Nat Med 2016. In a recent review (Abi-Dargham et al. World Psychiatry 2023), the FSA was highlighted as one of the most advanced biomarkers in schizophrenia, at the stage of external validation. B: Summary of the development and leave-one-site-out cross-validation of the FSA as a diagnostic biomarker, as in Li et al. Nat Med 2020. C: The current work consists in calculating FSA scores using the same method as in the original research to replicate the classification of diagnosis and correlation with treatment response by the FSA, to corroborate external validation.

## Methods

### Participant characteristics

We included data from the Human Connectome Project for Early Psychosis as in its 1.1 public release of August 2021^9^. Participants were patients within 5 years of having been diagnosed with either non-affective (schizophrenia, schizoaffective, schizophreniform, psychosis not otherwise specified) or affective psychosis (major depressive disorder with psychotic features and bipolar disorder with psychotic features) (total n=101, n=78 non-affective and n=23 affective psychosis), and healthy controls (n=51). All participants were administered the SCID-5^10^ diagnostic interview, which generated rule in and rule out diagnoses. Main entry criteria included age 16-35 and confirmation of primary psychosis for patients and ruling out history of primary psychosis or active affective, substance use, or anxiety disorder for controls. Patients were engaged in treatment with antipsychotics (mean dose in chlorpromazine equivalents = 104.3 mg) and overall, were mildly symptomatic (mean total PANSS = 46.29, SD=16.58). Data were acquired at four sites: Indiana University, Brigham and Women’s Hospital, McLean Hospital, and Massachusetts General Hospital. All procedures were approved by the Partners Healthcare Human Research Committee/IRB and comply with the regulations set forth by the Declaration of Helsinki.

In addition, for the prediction of treatment response we used data from the Zucker Hillside Hospital cohort (ZHH cohort) 97 patients at the time of their first treatment for psychosis (non-affective n=69, affective psychosis n=28) with minimal exposure to antipsychotics (43% treatment naïve, median exposure = 5 days). All participants were between 18 and 35 at the time of enrollment. Diagnostic eligibility was confirmed by the Structured Clinical Interview for DSM-5 (SCID-5)^10^. Patients underwent a standardized treatment protocol with risperidone or aripiprazole for 12 weeks, and regular clinical ratings. Patients underwent assessment by the Brief Psychopathology Rating Scale (BPRS-A)^11^ at baseline and weeks 2,4,6,8 and 12, and change in symptom severity between last assessment and baseline was calculated. In addition, we calculated treatment response, defined as two consecutive ratings of much or very much improved on the CGI, as well as a rating of ≤3 on four psychosis-related items of the BPRS-A^11^, resulting in n=52 (53.61%) responders (being the rest either non-responders or early exits). All participants were scanned at treatment onset at the North Shore University Hospital in Manhasset, NY, after providing written, informed consent under a protocol approved by the Institutional Review Board (IRB) of the Feinstein Institutes for Medical Research at Northwell Health. Participant characteristics are described in Table S1.

### fMRI acquisition

Resting state fMRI (rs-fMRI) scans were collected on 3T Siemens Prisma scanners using a multi-band accelerated echo-planar imaging (EPI) sequence described in detail in the Human Connectome Project^12^. For each study participant, a T1-weighted scan (TR = 2400 msec, TE = 2.22 msec, voxel size = 0.8 mm3, scan length = 6 min, 38 s) and two rs-fMRI sessions (TR=800ms, TE=37ms) were acquired at each timepoint. During each fMRI session, two runs of 5-min 47-s scans (i.e. 420 volumes) were collected in opposite phase encoding directions (one with PA and the other with AP) Rs-fMRI scans were collected with eyes open. To ensure signal stability, the first 13 volumes were discarded in data analysis. Each rs-fMRI scan consisted of 72 contiguous axial/oblique slices in the AC-PC orientation (TR = 720 ms, TE = 33.1 ms, matrix = 104 × 90, FOV = 208 mm, voxel = 2x2x2mm, multi-band acceleration factor = 8). The only major differences in acquisition for the scans from the ZHH cohort were that the rsMRI runs were of 7-min 17-s rsMRI runs, one each with AP and PA phase encoding directions (total 2 runs at each timepoint, 594 volumes each), and that images were acquired with eyes closed, with verification of wakefulness.

### Calculation of FSA scores and intrinsic connectivity of canonical networks

#### Image preprocessing

Scans from both datasets were preprocessed using HCP based pipelines^13^. Briefly, structural preprocessing included gradient distortion correction, brain extraction, cross-modal registration of T2 weighted (T2w) images to T1w, bias field correction based on square root (T1w*T2w) and non-linear registration to MNI space. The functional preprocessing methods used were gradient distortion correction, motion correction, and EPI image distortion correction based on spin-echo EPI field maps (FSL toolbox “topup”), and spatial registration to T1w image and MNI space^13^. An initial high pass filter of 2000 Hz was applied to remove slow drift trends before nuisance regression was performed using FMRIB’s ICA-based X-noiseifier (FIX)^14–16^.

Functional images then underwent 5-mm full-width-at-half-maximum spatial smoothing. Finally, we ran global signal regression (GSR), since this step was taken in the original FSA publication^5^, and also additional literature has suggested that GSR may facilitate behavioral predictions from rs-fMRI^17^. However, both GSR and no-GSR results are presented. To control for head motion, frame-wise displacement (FD) was calculated for each scan time point^18^. We applied a stringent motion threshold so that individuals for whom >30% of volumes had FD>0.3 were excluded from the analyses. This led to the exclusion of 13 and 8 individuals in the HCP for Early Psychosis dataset and ZHH cohort respectively.

#### Calculation of FSA and intrinsic connectivity of canonical networks

For the FSA calculation, we followed the procedures described in the original study^5^, using publicly available scripts for the calculation of the FSA^19^. The input to calculate the FSA were the preprocessed resting state fMRI images as described above, resliced to 3mm isotropic voxels, and fALFF images^20^ that we calculated separately using the RestPlus toolbox in Matlab^21^. For this step, we used preprocessed images as described above. Fourier transformation was used at every voxel to calculate the power of BOLD signal in the low frequency range of 0.01-0.08 Hz, and then divided by the entire frequency range. Both resting state and fALFF images were finally used to calculate the FSA scores, which reflect the distance to the separating hyperplane between cases and controls based on the model developed in the original publication^5^. For each scan, we generated FSA values for each run (i.e., 1 and 2), phase encoding direction (PA and AP) and GSR (with and without).

In addition to the FSA, we measured the intrinsic connectivity of canonical networks for each subject using custom python scripts based on nilearn^22^. Metrics from these networks served as a control for our analyses and also to put the FSA in perspective from well validated connectivity measures^8^. For this step, we generated voxel-wise connectivity matrices for each scan and fetched the ‘Dictionaries of Functional Modes’ (DiFuMo) atlas^23^ subsequently deriving functional connectivity for the Yeo networks^8^: cognitive control, default mode, dorsal attention, salience, somatosensory and visual. We extracted and averaged the correlations between nodes within each network as the intrinsic connectivity value for each subject.

#### Generation of values based on scans of increasing length

We used concatenation and slicing to generate scans of increasing length from which to calculate FSA and network intrinsic connectivity values. For this step, we normalized, mean centered, and concatenated the two consecutive runs in each phase encoding direction for each individual’s rs-fMRI scan (i.e., run 1 AP with run 2 AP etc…). Subsequently, we sliced each ∼10’ concatenated scan in 10 increments (i.e., first 82 volumes, first 164 volumes etc, up to the entire 820 volumes). Finally, we preprocessed each increment and analyzed it to obtain FSA and intrinsic network connectivity values as described above.

### Analyses

#### Distribution and potential confounding

We compared the distribution of the FSA and the network intrinsic connectivity by participant type (patients vs controls) and calculated the corresponding effect sizes of the difference in values between patients and controls. Then, we ran a linear regression including as dependent variable the FSA and independent variables age, sex, race, medication dose, and illness severity score to measure whether these variables may behave as confounding. In addition, we examined whether these variables affected differently patients or controls by running group interactions.

#### Classification performance and correlation with symptom improvement

We calculated receiver operating characteristic (ROC) curves to estimate the accuracy of classification of diagnosis. Analyses were repeated separately for patient sub-groups that only included affective and non-affective psychoses. Since the FSA was developed on patients with non-affective psychosis, we hypothesized that it would perform better in this population. We ran separate ROCs models for FSA derived from PA GSR, PA NoGSR, AP GSR and AP NoGSR scans. Classification accuracy was measured by the area under the curve (AUC) of the ROC curve, and 95% confidence intervals (95%CIs) were generated using 2000 bootstraps.

ROCs were also used to identify the score with the best discriminating ability (i.e., value with greatest true positive and lowest false positive fraction), which was then used to calculate the sensitivity and specificity for that discriminating threshold. As a reference, it has been suggested that AUC >80% is necessary for clinical utility of a biomarker^24^ although lower accuracy may still be useful depending on consequences of wrong classification and alternatives to the biomarker^4^.

To replicate the post-hoc finding reported in the original publication, or a correlation between baseline FSA score and symptom improvement over an antipsychotic trial, we ran a correlation between FSA scores derived for each phase encoding direction and GSR condition, and symptom improvement defined as difference in total BPRS-A score between the baseline assessment and the last available measure.

#### Test-retest and phase encoding direction reliability

We compared the FSA and intrinsic network connectivity from each run (i.e., test-retest reliability) and PA vs AP scans (i.e., phase encoding direction reliability) by using the intraclass correlation coefficient (ICC), an established measure of reliability^25^ that reflects the ratio of within-individual variance over total variance. Specifically, we used a two-way mixed, single score intraclass correlation coefficient [ICC(3,1)] generating also 95% CIs, which is recommended to test the reliability of the same measure by one rater^26^. We calculated reliabilities for the entire sample, as well as separately for patients and controls. As a rule of thumb ICC is deemed poor <0.4, fair 0.4–0.59, good 0.6–0.74, and excellent >0.75^27^. The reason to focus on phase encoding direction reliability, in addition to test-retest, is based on recent findings from our group about general differences in reliability in the connectome by phase encoding direction^28^.

#### Effects of scan length on classification performance and reliability

For this particular part of the analyses, we re-calculated FSA and network intrinsic connectivity values from the scans of increasing lengths described above. For values generated from scans of each duration, we calculated AUCs with corresponding 95%CIs, as well ICC with 95%CIs for phase encoding direction reliability.

Analyses were conducted with custom scripts in R and python available on https://github.com/lorente01

## RESULTS

### Distribution and potential confounding

FSA values were significantly lower in patients than in controls (p<0.0001) for both phase encoding directions and GSR analyses, in the same direction as in the original publication^5^, with effect sizes that ranged between cohen’s d=0.79 (95%CI=0.44-1.15) for PA No GSR to d=0.91 (95%CI=0.55-1.26) (Figure 2; Table S2). Differences between patients and controls were for the most part not significant for intrinsic network connectivity, and when significant of small effect size. Out of all the networks and scans (i.e., PA vs AP and GSR vs No GSR, only somatosensory network PA No GSR (d=0.41 (95%CI=0.07-0.75) and default mode network AP No GSR (d=0.35 (95%CI=0.01-0.68) were different between patients and controls. As expected, network intrinsic connectivity values were lower in scans that were subject to GSR. Age, sex, race, medication dose, and illness severity score did not have any significant impact on FSA values (Table S3).

**Figure 2.**
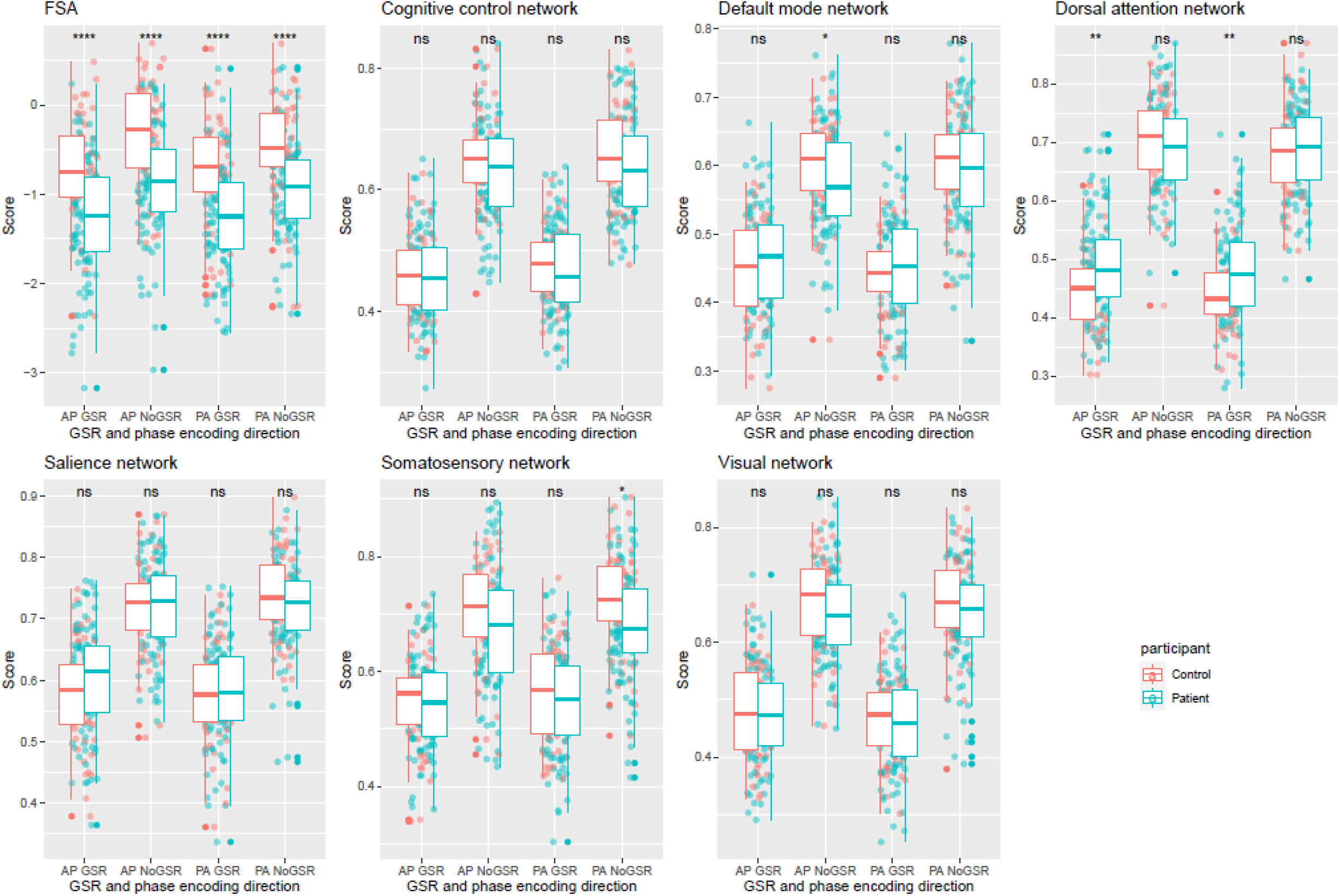
Distribution of FSA and network intrinsic connectivity values between patients and controls. **Footnote:** Distribution of FSA and intrinsic network connectivity values derived from scans in both phase encoding directions (posterior to anterior and anterior to posterior) with and without global signal regression by participant group (patients vs controls). **Legend:** AP: Anterior to posterior; FSA: Functional Striatal Abnormality score; GSR: Global signal regression; NoGSR: No global signal regression; ns: non-significant; PA: Posterior to anterior; ****: p Value <0.0001

### Classification performance and correlation with symptom improvement

For classification of diagnosis (i.e., psychosis vs healthy control), the classification performance of the FSA ranged between AUC=75.7%, (95%CI=67.3%-84.1%) for PA scans with GSR with sensitivity of 82% and specificity of 65%, and AUC=73.6%, (95%CI=65.0%-82.2%) for AP scans without GSR with sensitivity of 76% and specificity of 66%. When the discrimination was between non-affective psychosis and healthy controls, the classification ranged between AUC=80.5% (95%CI=72.1-88.0%) for PA scans with GSR with specificity of 82% and sensitivity of 73% and AUC=74.5% (95%CI=65.2-82.1%) for AP scans without GSR with specificity of 65% and specificity of 77%, whereas when the discrimination was between affective psychosis and healthy controls the classification ranged between AUC=70.2% (95%CI=57.4-81.9%) for AP scans without GSR with specificity of 43% and specificity of 96% and AUC=58.7% (95%CI=44.2-72.0%) for PA scans with GSR with sensitivity of 82% and specificity of 39% (Figure 3, panels A-C; Table S4).

**Figure 3.**
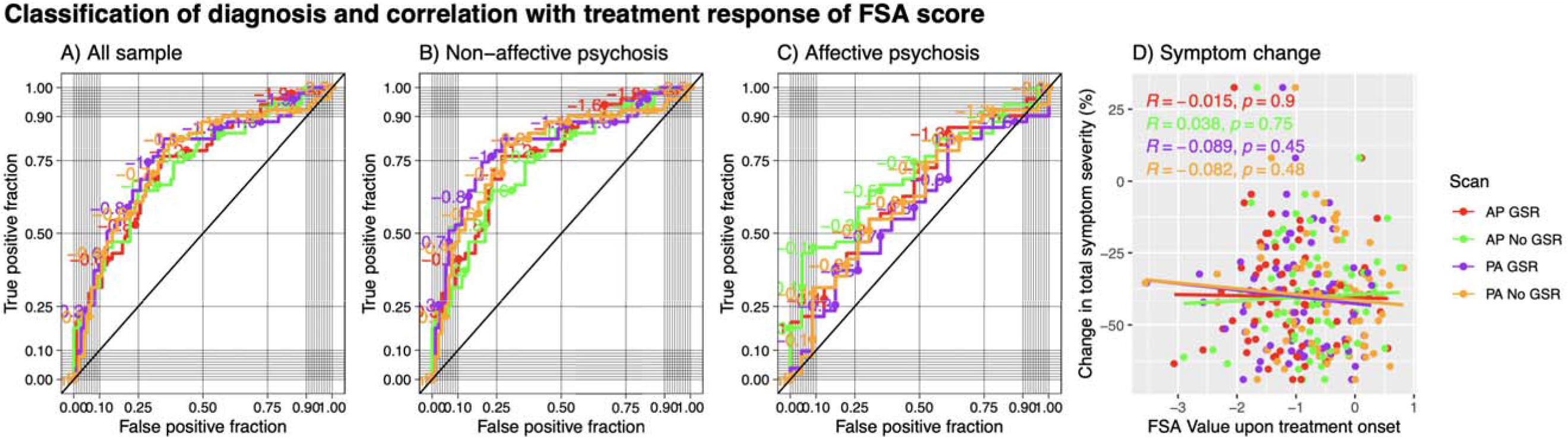
Receiver operating characteristic curves for prediction of diagnosis by FSA score and prediction of treatment response. **Footnote:** Classification of diagnosis and treatment response by FSA scores (Panels A to C) and correlation between baseline FSA scores. Each panel represents FSA scores derived from anterior to posterior (AP) and posterior to anterior (PA) phase encoding direction scans, with and without global signal regression. Diagonal line represents chance classification. Panel A represents all the sample, panel B represents the discrimination in diagnosis between non-affective psychosis and healthy controls, and panel C represents the discrimination in diagnosis between affective psychosis and healthy controls. Panel D represents % change in total symptom severity over a course of 12 weeks of antipsychotic treatment in individuals with acute psychosis, also with FSA scores derived from anterior to posterior (AP) and posterior to anterior (PA) phase encoding direction scans, with and without global signal regression. **Legend:** AP: Anterior to posterior; FSA: Functional Striatal Abnormality score; GSR: Global signal regression; NoGSR: No global signal regression; PA: Posterior to anterior.

When we tested correlation between baseline FSA scores and % change in total symptom severity in the ZHH cohort, we found that the regression coefficients ranged between r=0.038 p=0.75 for AP scans without GSR and r=0.089 p=0.45 for PA scans with GSR (Figure 3, panel D). When using a dichotomous definition of treatment response, classification performance ranged between AUC=55.7% (95%CI=40.4-70.8%) for PA scans with GSR with sensitivity of 35% and specificity of 88%, and AUC=49.3% (95%CI=34.4-65.9%) for AP scans with GSR with sensitivity of 72% and specificity of 41%. When the classification was between non-affective psychosis and healthy controls, the classification performance ranged between AUC=55.7% (95%CI=39.2-51.5%) for PA scans with GSR with sensitivity of 31% and specificity of 93%, and AUC=47.9% (95%CI=33.2-63.7%) for PA scans without GSR with sensitivity of 31% and specificity of 93%. There were not enough individuals with affective psychosis with enough data on treatment response to run separate classification analyses (Table S5; Figure S1).

### Test-retest and phase encoding direction reliability

Test-retest reliability between two runs within the same scan session for the FSA ranged between ICC=0.48 (95%CI=0.35-0.6) for AP scans without GSR, and ICC=0.22 (95%CI=0.06-0.37) for PA scans without GSR. Test-retest reliability for network intrinsic connectivity ranged between ICC=0.5 (95%CI=0.37-0.61) for cognitive control network in AP scans with GSR and ICC=0.07 (95%CI=-0.09-0.22) in salience network with PA No GSR scans (Figure 4, panel A; Table S6).

**Figure 4.**
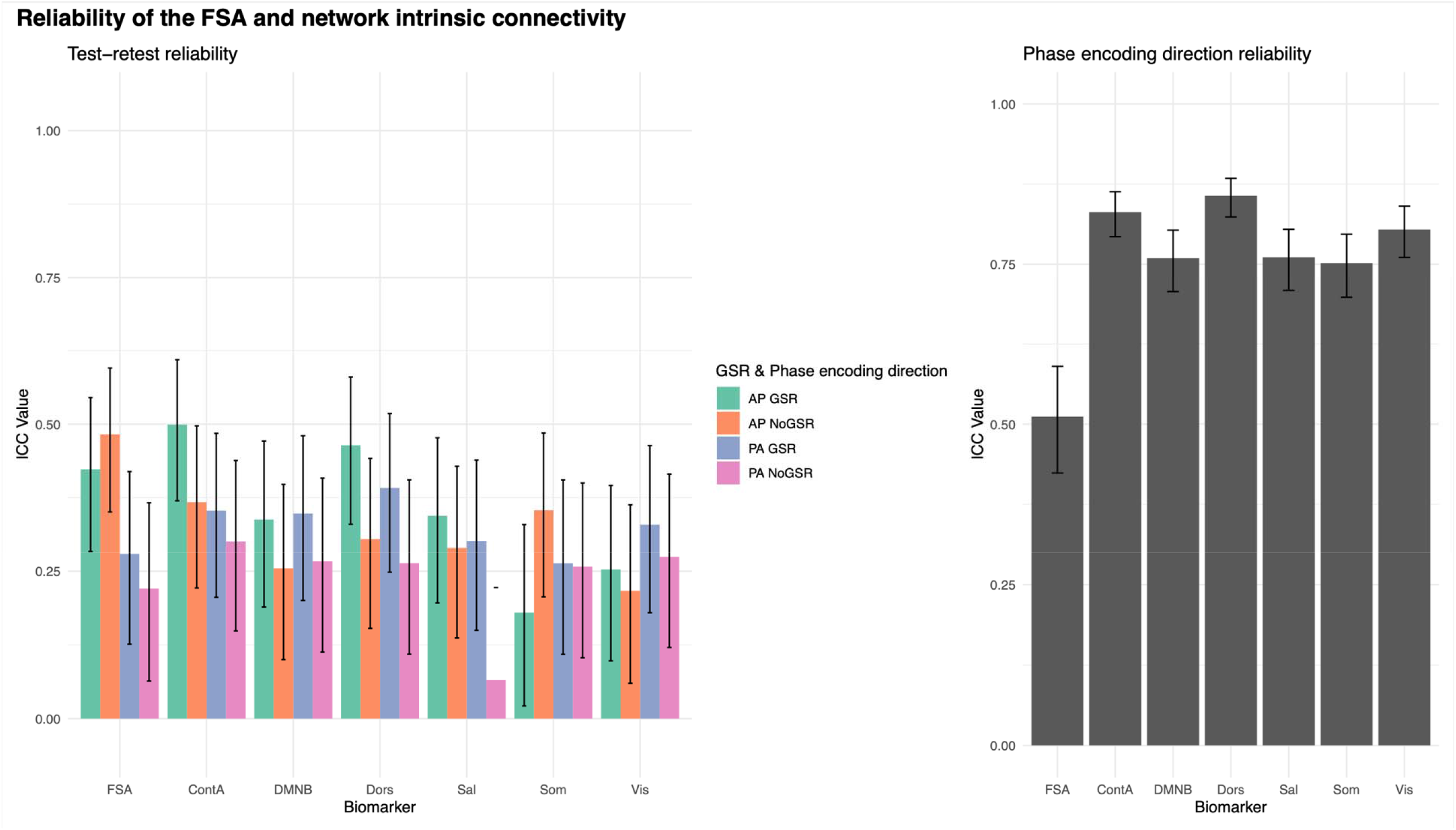
Test-retest and phase encoding direction reliability of the FSA and network intrinsic connectivity. **Footnote:** Test-retest (left) and phase encoding direction (right) reliability of FSA and intrinsic network connectivity. For test-retest reliability (left) each measure is derived respectively from anterior to posterior (AP) and posterior to anterior (PA) phase encoding direction scans, with and without global signal regression. In phase encoding direction reliability (right) averaged measures from two consecutive runs in the same phase encoding direction are being compared in scans with global signal regression. **Legend:** AP: Anterior to posterior; ContA: Intrinsic reliability in cognitive control network; DMNB: Intrinsic connectivity of default mode network; Dors: Intrinsic connectivity of dorsal attention network, FSA: Functional Striatal Abnormality score; GSR: Global signal regression; NoGSR: No global signal regression; PA: Posterior to anterior; Sal: Intrinsic connectivity of salience network, Som: intrinsic connectivity of somatosensory network; Vis: Intrinsic connectivity of visual network.

Phase encoding direction reliability for the FSA was ICC=0.51 (95%CI=0.42-0.59), while it ranged between ICC=0.86 (95%CI=0.82-0.88) for the dorsal attention network and ICC=0.75 (95%CI=0.70-0.80) for the somatosensory network (Figure 4, panel B; Table S7).

### Effects of scan length on classification performance and reliability

Using a slice of approximately 2’ (164 volumes) out of the concatenated runs (total 820 volumes) the diagnostic classification of the FSA ranged between AUC=67.2% (95%CI=57.6-76.7%) for AP scans without GSR and AUC=71.7% (95%CI=63.1-80.3%) for PA scans with GSR. When the entire concatenated scan was used, the diagnostic classification ranged between AUC=73.6% (95%CI=65.0-82.2%) for AP scans without GSR and AUC=75.7% (95%CI=67.3-84.1%) for PA scans with GSR. None of the classifications made by network intrinsic connectivity were significant, regardless of the scan time (Figure 5).

**Figure 5.**
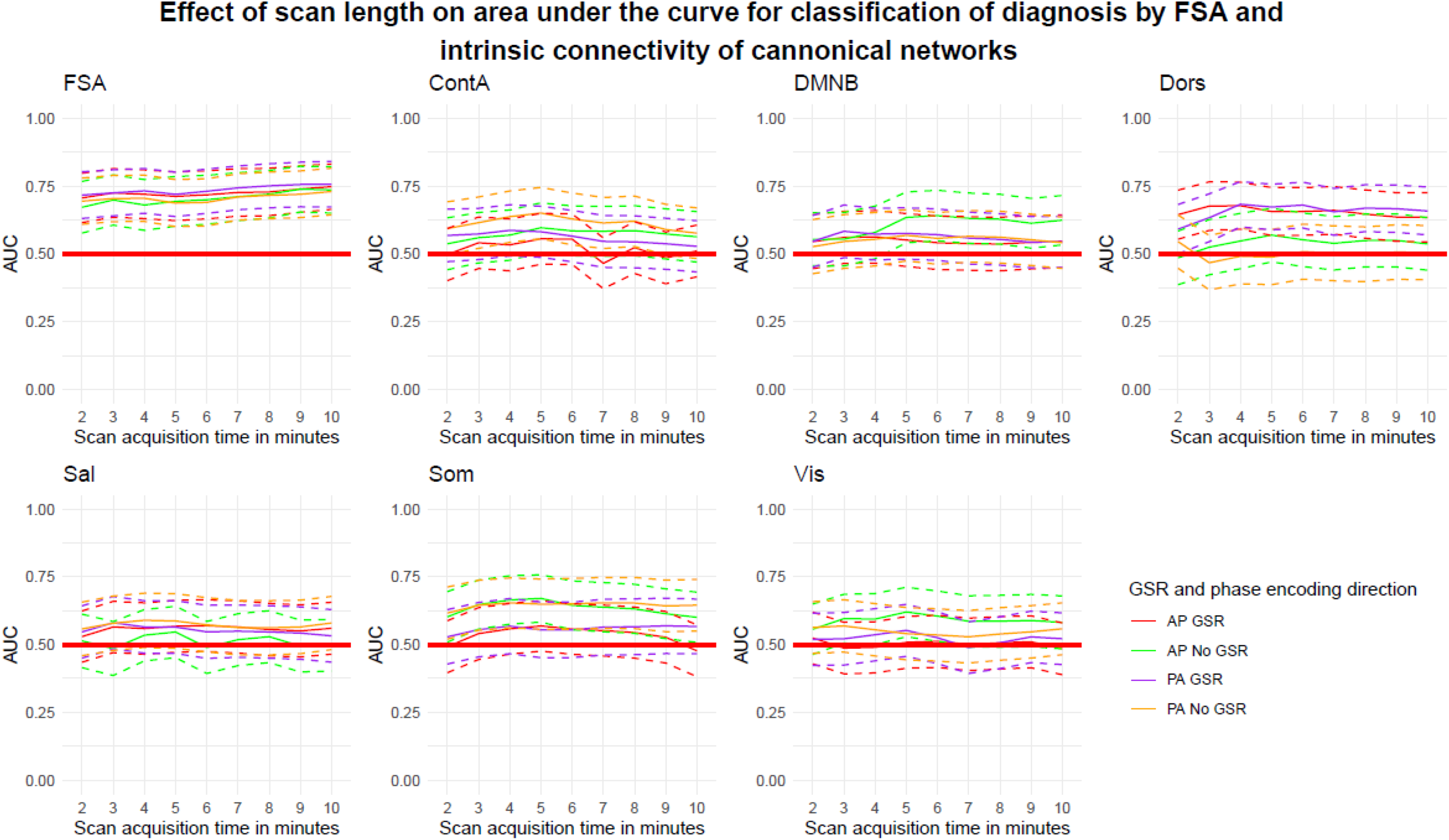
Accuracy of discrimination between cases and controls by scan acquisition time by the FSA and by intrinsic network connectivity. **Footnote:** Dotted lines represent 95%CI for AUC for increasing scan length. **Legend:** AP: Anterior to posterior; ContA: Intrinsic reliability in cognitive control network; DMNB: Intrinsic connectivity of default mode network; Dors: Intrinsic connectivity of dorsal attention network, FSA: Functional Striatal Abnormality score; GSR: Global signal regression; NoGSR: No global signal regression; PA: Posterior to anterior; Sal: Intrinsic connectivity of salience network, Som: intrinsic connectivity of somatosensory network; Vis: Intrinsic connectivity of visual network.

When the same slice scheme was used to test phase encoding direction reliability, we found significant increments in reliability for the FSA, starting from ICC=0.29 (95%CI=0.14-0.43) in slices of approximately 2’ duration to ICC=0.56 (95%CI=0.45-0.66) when using the entire concatenated scan. Similar increments were observed across network intrinsic connectivity values (Figure 6).

**Figure 6.**
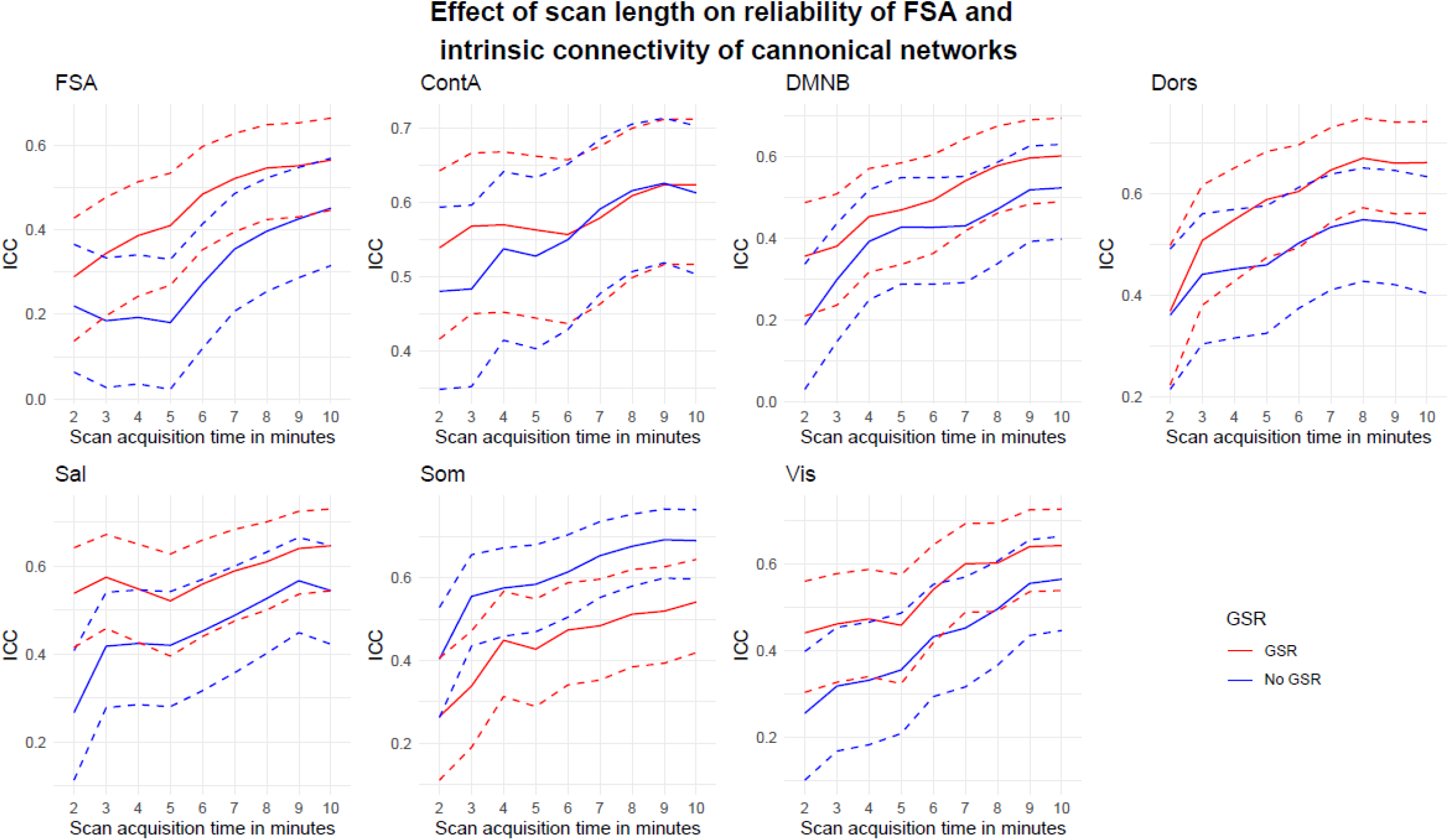
Phase encoding direction reliability by scan acquisition time for the FSA and by intrinsic network connectivity. **Footnote:** Dotted lines represent 95%CI for ICC of phase encoding direction reliability for increasing scan length. **Legend:** AP: Anterior to posterior; ContA: Intrinsic reliability in cognitive control network; DMNB: Intrinsic connectivity of default mode network; Dors: Intrinsic connectivity of dorsal attention network, FSA: Functional Striatal Abnormality score; GSR: Global signal regression; NoGSR: No global signal regression; PA: Posterior to anterior; Sal: Intrinsic connectivity of salience network, Som: intrinsic connectivity of somatosensory network; Vis: Intrinsic connectivity of visual network.

## DISCUSSION

Using a public independent dataset, we found that the original findings on the performance of the FSA as a diagnostic biomarker for schizophrenia^5^ are generalizable out of sample. However, the post-hoc finding of the association between treatment response and baseline FSA scores in the original report^5^ did not replicate in a separate treatment response dataset, suggesting that while as a diagnostic biomarker of schizophrenia the FSA has demonstrated external validity, it may be limited as a prognostic biomarker in its current form. Furthermore, this biomarker showed test-retest reliability comparable -and in some cases superior – to that of the Yeo networks, which have been previously well-validated^8^. Like recent work by our group ^28^, we identified differences in reliability by phase encoding direction, which seemed to be more pronounced in the FSA than in network intrinsic connectivity, although this did not have any significant impact in the accuracy of predictions. Similarly, there seemed to be greater gains in reliability than in accuracy of predictions by increasing the scan length.

Despite differences in participant demographic and clinical characteristics, scanner type, acquisition parameters, and preprocessing approach – all of which may reduce the reliability of fMRI scans and potentially the reproducibility of findings^29–31^ – the results in the Human Connectome Project are quite consistent with those of the Chinese datasets in which it was developed and initially tested. In addition to a classification accuracy of 75.7% (vs 80.1% in the original publication), we did also observe consistently higher FSA values in controls than in patients, improvement in classification accuracy when only non-affective psychoses were included, and better results with global signal regression, all of which were reported in the original publication^5^. Furthermore, we did not find significant effects of demographic, clinical, or acquisition variables on FSA values, highlighting the internal validity of this biomarker. The replication of these findings attests to the value of the approach used to develop the FSA: advanced multivariable classification methods on large datasets with leave-one-site-out cross-validation. Similar methods applying data-hungry models with contingencies to mitigate overfitting have been successfully applied to other areas of medicine, such as liquid biopsy for cancer^32^, radiomics in medical imaging^33^, or ophthalmology^34^. While diagnostic biomarkers have limited applicability in psychiatric conditions – the psychiatric interview is still necessary to assess the needs of each individual in addition to making a diagnosis – these results bring closer the application of neuroimaging biomarkers in psychiatric disorders as a test case.

An application more likely to be adopted in the clinic is that of prognostic biomarker. Predicting that an individual will not respond to an antipsychotic could avoid unsatisfactory experiences with ineffective treatments, a known risk factor of non-adherence^35^ and ultimately relapse^36^. Furthermore, it could reduce the time while someone is acutely psychotic – and potentially a danger to self or others – by allowing the expedited use of clozapine, the only drug approved for treatment resistant schizophrenia^37^. Neuroimaging biomarkers could be very helpful for this purpose since clinical information alone is rather limited to predict treatment response^38^. However, it may be necessary to develop these biomarkers using datasets from treatment response, instead of datasets of cases vs controls. It is not entirely surprising that despite robust performance for diagnosis classification, the FSA – for which features were selected using case-control datasets – was limited as a prognostic biomarker. Although difficult to compare, meta-analyses on functional connectivity in schizophrenia vs controls^39^, and in treatment responders vs non-responders^40^ do not necessarily overlap in their findings, aligning with the idea that the pathophysiology of the schizophrenia syndrome may go beyond the mechanisms engaged by antipsychotic drugs. In fact, in about one third of individuals with schizophrenia, positive symptoms fail to respond to antipsychotic drugs^41^, and furthermore negative symptoms or cognitive deficits, core aspects of the syndrome, are not engaged by antipsychotic medication^42^. Thus, the mechanisms involved in treatment response likely account for only a proportion of the connectivity abnormalities observed in schizophrenia that were used for feature selection of the FSA. A similar example is the application of polygenic risk scores (PRS) in schizophrenia. PRSs can classify individuals with schizophrenia with AUCs ranging between 60 and 66%^43^, but in general the proportion of the variance on treatment response explained by schizophrenia PRSs is lower^44^. Overall, this line of research suggests that applying approaches like the ones used to develop the FSA on large datasets of treatment response could be helpful to optimize prognostic biomarkers in schizophrenia and bring them closer to clinical application.

We observed effects of the phase encoding direction of the scan on reliability. Compared with network intrinsic network connectivity, the reliability of FSA values generated from PA vs AP scans was significantly lower, suggesting that FSA values are more vulnerable to phase encoding direction effects than other connectivity measures. AP scans showed better test-retest reliability; however, the predictions of AP and PA scans were not significantly different, suggesting that phase encoding direction may be more relevant for reliability than for accuracy of predictions. Similarly, we observed significant improvements in phase encoding direction reliability by increasing the acquisition length, however this had little effect on predictions, which were already good with scans of about 2 minutes. This apparent decoupling between accuracy of predictions and reliability has indeed been well documented ^45–48^, and indeed reflects the fact that accuracy and reliability are related but independent constructs. For instance, a given measure may get better reliability by capitalizing on reliable but uninformative features in the connectome (i.e., consistent noise), whereas other measures may capitalize on various states that while different, are all related to the phenotype of interest. Thus, it has been argued that since these discrepancies may occur, the focus in biomarker development should be on accuracy of predictions rather than on reliability^48^. This being said, to our knowledge in the development of the FSA there was no explicit management of the potential effects of phase encoding direction. These data indicate that in future iterations of biomarker development at the very least it should be explicitly managed in the analyses, and ideally at the stage of image acquisition.

These data should be interpreted in the light of several limitations. First, the sample size in this replication was relatively small (n=152) compared to the development sample size in the original study (n=1,100). However, this sample size is comparable to the sample sizes for the cross-validation sets that were used in the original analyses. Second, we did not test the specificity of the replication by attempting to measure the discrimination of diagnoses other than psychosis, although we observed that predictions were significantly better for discrimination between non-affective psychosis and healthy controls than between affective psychosis and healthy controls. Third, treatment response was tested on a cohort slightly different from the cohorts used to test treatment response in the original FSA study (multiepisode patients treated with both clozapine and non-clozapine antipsychotics vs first episode patients treated with non-clozapine antipsychotics). Fourth, test-retest reliability could only be tested for the individual runs, but only reliability between scans of different phase encoding direction (i.e., PA vs AP) could be tested used in the time analysis since we concatenated both runs to obtain longer scans to test the effects of acquisition length. Fifth, despite concatenation of both runs, scans were relatively short and did not reach a plateau in the time analysis, thus not allowing to conclude about the minimal scan duration for best reliability and accuracy.

In conclusion, using the framework for biomarker development of consecutive contingent phases – target identification, internal validity, external validity, and demonstration of clinical utility^3^ – this work emphasizes the internal validity of the FSA given the limited effect of common confounding, such as demographic or clinical characteristics. It also confirms the external validity as a diagnostic biomarker by replicating the original findings out of sample under rather different conditions. By and large, the test-retest reliability of the FSA comparable to that of the intrinsic connectivity in the Yeo networks^8^, which have been well studied. These findings demonstrate the value of multi-site research to develop large datasets in which data hungry advanced statistical techniques can be applied, yielding robust results. To move forward biomarker development in schizophrenia – from demonstrating external validity to clinical utility – it is necessary to apply similar approaches to treatment response data, to generate robust prognostic biomarkers that can predict treatment response.

## Data Availability

Data for the Human Connectome Project for Early Psychosis are public. Analyses were conducted with custom scripts in R and python available on https://github.com/lorente01

## Acknowledgements

JR was supported by NIH grant K23MH127300.

## Supplementary information

Supplementary information is available at MP’s website

