## Supplementary material for "Replication of a neuroimaging biomarker for striatal dysfunction in psychosis"

**Supplementary Information**

Table S1. Sample characteristics

| **HCP psychosis dataset (n=152)** | | | **ZHH treatment response cohort (n=97)** | |
| --- | --- | --- | --- | --- |
|  | Early phase psychosis | Healthy controls |  | First episode psychosis |
| N | n= 101 | n= 51 |  | n= 97 |
| Female (n,%) | n=38, (37.62%) | n=19, (37.25%) | Female (n,%) | n=44, (45.36%) |
| Age (mean,sd) | 22.49, (3.54) | 24.83, (4.23) | Age (mean,sd) | 24.3, (5.9) |
| White (n,%) | n=52, (51.49%) | n=37, (72.55%) | White (n,%) | n=32, (32.99%) |

Table S2. Effect sizes of differences between patients and controls in FSA and intrinsic connectivity values of canonical networks

| Connectivity measure | Phase encoding and GSR | Cohens d | lower bound 95%CI | upper bound 95%CI |
| --- | --- | --- | --- | --- |
| FSA | AP GSR | 0.88 | 0.53 | 1.23 |
| FSA | AP No GSR | 0.86 | 0.51 | 1.21 |
| FSA | PA GSR | 0.91 | 0.56 | 1.26 |
| FSA | PA No GSR | 0.80 | 0.45 | 1.15 |
| Cognitive control network | AP GSR | 0.01 | -0.32 | 0.35 |
| Cognitive control network | AP No GSR | 0.24 | -0.10 | 0.58 |
| Cognitive control network | PA GSR | 0.11 | -0.22 | 0.45 |
| Cognitive control network | PA No GSR | 0.25 | -0.09 | 0.58 |
| Default mode network | AP GSR | -0.25 | -0.59 | 0.09 |
| Default mode network | AP No GSR | 0.35 | 0.01 | 0.68 |
| Default mode network | PA GSR | -0.19 | -0.52 | 0.15 |
| Default mode network | PA No GSR | 0.10 | -0.23 | 0.44 |
| Dorsal attention network | AP GSR | -0.52 | -0.86 | -0.18 |
| Dorsal attention network | AP No GSR | 0.06 | -0.28 | 0.39 |
| Dorsal attention network | PA GSR | -0.48 | -0.82 | -0.14 |
| Dorsal attention network | PA No GSR | -0.01 | -0.34 | 0.33 |
| Salience network | AP GSR | -0.28 | -0.62 | 0.06 |
| Salience network | AP No GSR | -0.05 | -0.39 | 0.29 |
| Salience network | PA GSR | -0.11 | -0.45 | 0.23 |
| Salience network | PA No GSR | 0.25 | -0.09 | 0.59 |
| Somatosensory network | AP GSR | 0.03 | -0.31 | 0.37 |
| Somatosensory network | AP No GSR | 0.28 | -0.06 | 0.62 |
| Somatosensory network | PA GSR | 0.16 | -0.18 | 0.49 |
| Somatosensory network | PA No GSR | 0.41 | 0.07 | 0.75 |
| Visual network | AP GSR | 0.03 | -0.31 | 0.36 |
| Visual network | AP No GSR | 0.18 | -0.16 | 0.52 |
| Visual network | PA GSR | 0.12 | -0.22 | 0.45 |
| Visual network | PA No GSR | 0.22 | -0.11 | 0.56 |

Table S3. Effect of potential confounders on FSA (p Values)

| Measure | p Value Age | p Value Sex | p Value Race | p Value mean FD | p Value Age*Group | p Value Sex*Group | p Value Race*Group | p Value mean FD*Group | p Value PANSS (SZ only) | p Value CPZ eq (SZ only) |
| --- | --- | --- | --- | --- | --- | --- | --- | --- | --- | --- |
| FSA PA GSR | 0.72 | 0.69 | 0.07 | 0.88 | 0.13 | 0.49 | 0.23 | 0.26 | 0.27 | 0.23 |
| FSA PA No GSR | 0.44 | 0.09 | 0.01 | 0.49 | 0.18 | 0.83 | 0.13 | 0.44 | 0.34 | 0.06 |
| FSA AP GSR | 0.97 | 0.86 | 0.74 | 0.72 | 0.66 | 0.25 | 0.50 | 0.90 | 0.80 | 0.06 |
| FSA AP No GSR | 0.64 | 0.73 | 0.85 | 0.19 | 0.44 | 0.40 | 0.81 | 0.35 | 0.79 | 0.13 |

Table S4. Predictive performance of the FSA discriminating individuals with psychosis from healthy controls

| **Entire cohort** | | | | | | |
| --- | --- | --- | --- | --- | --- | --- |
| FSA measure | Sensitivity | Specificity | Optimal discrimination cutoff | AUC | Lower bound 95%CI | Upper bound 95%CI |
| FSA AP GSR | 0.76 | 0.66 | -1.04 | 0.74 | 0.65 | 0.81 |
| FSA AP No GSR | 0.65 | 0.75 | -0.45 | 0.73 | 0.64 | 0.81 |
| FSA PA GSR | 0.82 | 0.65 | -1.06 | 0.75 | 0.67 | 0.83 |
| FSA PA No GSR | 0.78 | 0.65 | -0.73 | 0.75 | 0.66 | 0.83 |
| **Non-affective psychosis as cases only** | | | | | | |
| FSA measure | Sensitivity | Specificity | Optimal discrimination cutoff | AUC | Lower bound 95%CI | Upper bound 95%CI |
| FSA AP GSR | 0.76 | 0.73 | -1.04 | 0.76 | 0.68 | 0.85 |
| FSA AP No GSR | 0.65 | 0.77 | -0.45 | 0.74 | 0.65 | 0.82 |
| FSA PA GSR | 0.82 | 0.73 | -1.06 | 0.80 | 0.72 | 0.88 |
| FSA PA No GSR | 0.80 | 0.72 | -0.74 | 0.78 | 0.69 | 0.86 |
| **Affective psychosis as cases only** | | | | | | |
| FSA measure | Sensitivity | Specificity | Optimal discrimination cutoff | AUC | Lower bound 95%CI | Upper bound 95%CI |
| FSA AP GSR | 0.86 | 0.39 | -1.37 | 0.65 | 0.52 | 0.78 |
| FSA AP No GSR | 0.43 | 0.96 | -0.08 | 0.70 | 0.57 | 0.82 |
| FSA PA GSR | 0.82 | 0.39 | -1.06 | 0.59 | 0.45 | 0.73 |
| FSA PA No GSR | 0.55 | 0.70 | -0.52 | 0.63 | 0.49 | 0.76 |

Table S5. Predictive performance of the FSA discriminating in individuals with psychosis between response and non-response to a prospective trial of antipsychotic drugs

| **Entire cohort** | | | | | | |
| --- | --- | --- | --- | --- | --- | --- |
| FSA measure | Sensitivity | Specificity | Optimal discrimination cutoff | AUC | Lower bound 95%CI | Upper bound 95%CI |
| FSA AP GSR | 0.72 | 0.41 | -1.47 | 0.49 | 0.34 | 0.66 |
| FSA AP No GSR | 0.72 | 0.35 | -0.98 | 0.52 | 0.36 | 0.68 |
| FSA PA GSR | 0.35 | 0.88 | -0.54 | 0.56 | 0.40 | 0.71 |
| FSA PA No GSR | 0.35 | 0.88 | -0.02 | 0.53 | 0.38 | 0.69 |
| **Non-affective psychosis only** | | | | | | |
| FSA measure | Sensitivity | Specificity | Optimal discrimination cutoff | AUC | Lower bound 95%CI | Upper bound 95%CI |
| FSA AP GSR | 0.67 | 0.50 | -1.47 | 0.52 | 0.34 | 0.70 |
| FSA AP No GSR | 0.42 | 0.71 | -0.64 | 0.50 | 0.32 | 0.67 |
| FSA PA GSR | 0.31 | 0.93 | -0.54 | 0.56 | 0.39 | 0.71 |
| FSA PA No GSR | 0.31 | 0.93 | -0.02 | 0.48 | 0.33 | 0.64 |

Figure S1. Receiver operating characteristic curves for prediction of treatment response by FSA score at baseline

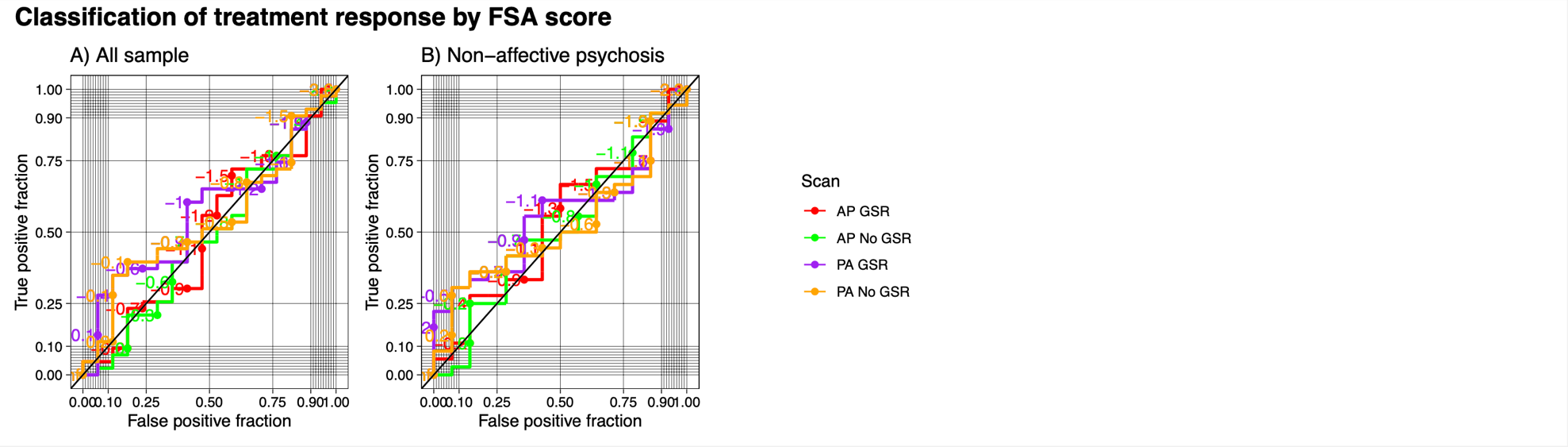

Table S6. Intraclass correlation coefficients of test-restest reliability of FSA and network intrinsic connectivity

| **Biomarker** | **GSR_phase** | **Participant type** | **ICC** | **lower bound 95%CI** | **upper bound 95%CI** |
| --- | --- | --- | --- | --- | --- |
| FSA | AP GSR | Entire sample | 0.42 | 0.28 | 0.55 |
| FSA | AP GSR | Control | 0.24 | -0.04 | 0.48 |
| FSA | AP GSR | Patient | 0.42 | 0.24 | 0.57 |
| FSA | AP NoGSR | Entire sample | 0.48 | 0.35 | 0.60 |
| FSA | AP NoGSR | Control | 0.46 | 0.21 | 0.65 |
| FSA | AP NoGSR | Patient | 0.41 | 0.23 | 0.56 |
| FSA | PA GSR | Entire sample | 0.28 | 0.13 | 0.42 |
| FSA | PA GSR | Control | 0.20 | -0.08 | 0.45 |
| FSA | PA GSR | Patient | 0.20 | 0.01 | 0.38 |
| FSA | PA NoGSR | Entire sample | 0.22 | 0.06 | 0.37 |
| FSA | PA NoGSR | Control | 0.28 | 0.00 | 0.51 |
| FSA | PA NoGSR | Patient | 0.10 | -0.10 | 0.29 |
| Cognitive control network | AP GSR | Entire sample | 0.50 | 0.37 | 0.61 |
| Cognitive control network | AP GSR | Control | 0.47 | 0.23 | 0.66 |
| Cognitive control network | AP GSR | Patient | 0.53 | 0.37 | 0.65 |
| Cognitive control network | AP NoGSR | Entire sample | 0.37 | 0.22 | 0.50 |
| Cognitive control network | AP NoGSR | Control | 0.34 | 0.07 | 0.56 |
| Cognitive control network | AP NoGSR | Patient | 0.39 | 0.21 | 0.54 |
| Cognitive control network | PA GSR | Entire sample | 0.35 | 0.21 | 0.48 |
| Cognitive control network | PA GSR | Control | 0.25 | -0.03 | 0.49 |
| Cognitive control network | PA GSR | Patient | 0.40 | 0.22 | 0.55 |
| Cognitive control network | PA NoGSR | Entire sample | 0.30 | 0.15 | 0.44 |
| Cognitive control network | PA NoGSR | Control | 0.23 | -0.05 | 0.47 |
| Cognitive control network | PA NoGSR | Patient | 0.35 | 0.16 | 0.51 |
| Default mode network | AP GSR | Entire sample | 0.34 | 0.19 | 0.47 |
| Default mode network | AP GSR | Control | 0.36 | 0.09 | 0.57 |
| Default mode network | AP GSR | Patient | 0.32 | 0.14 | 0.49 |
| Default mode network | AP NoGSR | Entire sample | 0.25 | 0.10 | 0.40 |
| Default mode network | AP NoGSR | Control | 0.14 | -0.13 | 0.40 |
| Default mode network | AP NoGSR | Patient | 0.30 | 0.11 | 0.46 |
| Default mode network | PA GSR | Entire sample | 0.35 | 0.20 | 0.48 |
| Default mode network | PA GSR | Control | 0.11 | -0.17 | 0.37 |
| Default mode network | PA GSR | Patient | 0.45 | 0.28 | 0.59 |
| Default mode network | PA NoGSR | Entire sample | 0.27 | 0.11 | 0.41 |
| Default mode network | PA NoGSR | Control | 0.13 | -0.15 | 0.39 |
| Default mode network | PA NoGSR | Patient | 0.32 | 0.13 | 0.48 |
| Dorsal attention network | AP GSR | Entire sample | 0.46 | 0.33 | 0.58 |
| Dorsal attention network | AP GSR | Control | 0.35 | 0.08 | 0.57 |
| Dorsal attention network | AP GSR | Patient | 0.48 | 0.31 | 0.61 |
| Dorsal attention network | AP NoGSR | Entire sample | 0.30 | 0.15 | 0.44 |
| Dorsal attention network | AP NoGSR | Control | 0.26 | -0.01 | 0.50 |
| Dorsal attention network | AP NoGSR | Patient | 0.33 | 0.15 | 0.50 |
| Dorsal attention network | PA GSR | Entire sample | 0.39 | 0.25 | 0.52 |
| Dorsal attention network | PA GSR | Control | 0.23 | -0.05 | 0.47 |
| Dorsal attention network | PA GSR | Patient | 0.44 | 0.26 | 0.58 |
| Dorsal attention network | PA NoGSR | Entire sample | 0.26 | 0.11 | 0.41 |
| Dorsal attention network | PA NoGSR | Control | 0.23 | -0.05 | 0.47 |
| Dorsal attention network | PA NoGSR | Patient | 0.29 | 0.10 | 0.46 |
| Salience network | AP GSR | Entire sample | 0.34 | 0.20 | 0.48 |
| Salience network | AP GSR | Control | 0.26 | -0.02 | 0.50 |
| Salience network | AP GSR | Patient | 0.38 | 0.20 | 0.54 |
| Salience network | AP NoGSR | Entire sample | 0.29 | 0.14 | 0.43 |
| Salience network | AP NoGSR | Control | 0.29 | 0.02 | 0.52 |
| Salience network | AP NoGSR | Patient | 0.29 | 0.11 | 0.46 |
| Salience network | PA GSR | Entire sample | 0.30 | 0.15 | 0.44 |
| Salience network | PA GSR | Control | 0.40 | 0.14 | 0.61 |
| Salience network | PA GSR | Patient | 0.27 | 0.08 | 0.44 |
| Salience network | PA NoGSR | Entire sample | 0.07 | -0.09 | 0.22 |
| Salience network | PA NoGSR | Control | 0.00 | -0.27 | 0.27 |
| Salience network | PA NoGSR | Patient | 0.10 | -0.10 | 0.29 |
| Somatosensory network | AP GSR | Entire sample | 0.18 | 0.02 | 0.33 |
| Somatosensory network | AP GSR | Control | 0.20 | -0.08 | 0.45 |
| Somatosensory network | AP GSR | Patient | 0.18 | -0.02 | 0.36 |
| Somatosensory network | AP NoGSR | Entire sample | 0.35 | 0.21 | 0.49 |
| Somatosensory network | AP NoGSR | Control | 0.16 | -0.12 | 0.42 |
| Somatosensory network | AP NoGSR | Patient | 0.48 | 0.32 | 0.62 |
| Somatosensory network | PA GSR | Entire sample | 0.26 | 0.11 | 0.41 |
| Somatosensory network | PA GSR | Control | 0.15 | -0.13 | 0.41 |
| Somatosensory network | PA GSR | Patient | 0.33 | 0.14 | 0.49 |
| Somatosensory network | PA NoGSR | Entire sample | 0.26 | 0.10 | 0.40 |
| Somatosensory network | PA NoGSR | Control | 0.17 | -0.10 | 0.43 |
| Somatosensory network | PA NoGSR | Patient | 0.29 | 0.10 | 0.46 |
| Visual network | AP GSR | Entire sample | 0.25 | 0.10 | 0.40 |
| Visual network | AP GSR | Control | 0.20 | -0.08 | 0.45 |
| Visual network | AP GSR | Patient | 0.28 | 0.09 | 0.45 |
| Visual network | AP NoGSR | Entire sample | 0.22 | 0.06 | 0.36 |
| Visual network | AP NoGSR | Control | 0.25 | -0.03 | 0.49 |
| Visual network | AP NoGSR | Patient | 0.22 | 0.02 | 0.40 |
| Visual network | PA GSR | Entire sample | 0.33 | 0.18 | 0.46 |
| Visual network | PA GSR | Control | 0.25 | -0.02 | 0.49 |
| Visual network | PA GSR | Patient | 0.38 | 0.20 | 0.53 |
| Visual network | PA NoGSR | Entire sample | 0.27 | 0.12 | 0.41 |
| Visual network | PA NoGSR | Control | 0.31 | 0.04 | 0.54 |
| Visual network | PA NoGSR | Patient | 0.26 | 0.07 | 0.43 |

Table S7. Intraclass correlation coefficients of phase encoding direction reliability (i.e., AP vs PA) of FSA and network intrinsic connectivity

| **Biomarker** | **Participant type** | **ICC** | **lower bound 95%CI** | **upper bound 95%CI** |
| --- | --- | --- | --- | --- |
| FSA | Entire sample | 0.51 | 0.42 | 0.59 |
| FSA | Patient | 0.54 | 0.43 | 0.63 |
| FSA | Control | 0.23 | 0.04 | 0.40 |
| Cognitive control network | Entire sample | 0.83 | 0.79 | 0.86 |
| Cognitive control network | Patient | 0.84 | 0.80 | 0.88 |
| Cognitive control network | Control | 0.81 | 0.73 | 0.86 |
| Default mode network | Entire sample | 0.76 | 0.71 | 0.80 |
| Default mode network | Patient | 0.75 | 0.68 | 0.80 |
| Default mode network | Control | 0.79 | 0.70 | 0.85 |
| Dorsal attention network | Entire sample | 0.86 | 0.82 | 0.88 |
| Dorsal attention network | Patient | 0.85 | 0.81 | 0.89 |
| Dorsal attention network | Control | 0.86 | 0.80 | 0.91 |
| Salience network | Entire sample | 0.76 | 0.71 | 0.80 |
| Salience network | Patient | 0.73 | 0.66 | 0.79 |
| Salience network | Control | 0.82 | 0.75 | 0.88 |
| Somatosensory network | Entire sample | 0.75 | 0.70 | 0.80 |
| Somatosensory network | Patient | 0.75 | 0.68 | 0.80 |
| Somatosensory network | Control | 0.76 | 0.66 | 0.83 |
| Visual network | Entire sample | 0.80 | 0.76 | 0.84 |
| Visual network | Patient | 0.83 | 0.78 | 0.87 |
| Visual network | Control | 0.76 | 0.66 | 0.83 |
